# Algorithmic Fairness and Bias Mitigation for Clinical Machine Learning: Insights from Rapid COVID-19 Diagnosis by Adversarial Learning

**DOI:** 10.1101/2022.01.13.22268948

**Authors:** Jenny Yang, Andrew A. S. Soltan, Yang Yang, David A. Clifton

## Abstract

Machine learning is becoming increasingly prominent in healthcare. Although its benefits are clear, growing attention is being given to how machine learning may exacerbate existing biases and disparities. In this study, we introduce an adversarial training framework that is capable of mitigating biases that may have been acquired through data collection or magnified during model development. For example, if one class is over-presented or errors/inconsistencies in practice are reflected in the training data, then a model can be biased by these. To evaluate our adversarial training framework, we used the statistical definition of equalized odds. We evaluated our model for the task of rapidly predicting COVID-19 for patients presenting to hospital emergency departments, and aimed to mitigate regional (hospital) and ethnic biases present. We trained our framework on a large, real-world COVID-19 dataset and demonstrated that adversarial training demonstrably improves outcome fairness (with respect to equalized odds), while still achieving clinically-effective screening performances (NPV*>*0.98). We compared our method to the benchmark set by related previous work, and performed prospective and external validation on four independent hospital cohorts. Our method can be generalized to any outcomes, models, and definitions of fairness.

## I. Introduction

### A. Background

A fundamental observation in machine learning research is that models are susceptible to biases present in training data, and that these biases can lead to poorer model outcomes and unfair decision-making. Here, an “unfair” decision refers to any outcome that is skewed towards a particular group or population [1]. This is particularly harmful in sensitive domains such as healthcare, as it not only leads to inaccurate predictions, but also propagates existing healthcare inequities and may compromise patient trust.

If a machine learning model acquires unintentional biases, it may be unable to capture the true relationship between the features and the target outcome, resulting in poorer model performance. Thus, bias mitigation can help a model generalize better across different populations and groups, resulting in a stronger classifier.

In addition to training a stronger classifier, bias mitigation is also relevant for the purpose of model fairness. As achieving generalizability often requires large datasets, many health-related projects may integrate datasets from multiple hospitals or institutions in order to increase the amount of data available for training. However, even if each respective dataset contains the same features, there is vast literature discussing minor and substantial disparities in health and healthcare practice across different geographic regions. Namely, disease prevalence/mortality, quality of healthcare services, and specific devices used (e.g. blood analysis devices) varies widely across hospitals in different regions. This heterogeneity has been acknowledged worldwide and has been examined for a range of medical conditions and diseases [2]-[4], as well as different drivers of healthcare quality [2], [5]. If these types of biases become reflected in a model’s decisions, then certain hospitals could be unintentionally isolated for exhibiting poorer outcomes, further widening interregional and interhospital inequality gaps, and also adversely affect model performance.

Health inequalities related to demographic biases such as sex, gender, age, and ethnicity, can also exist. For example, in terms of gender bias, physicians have been found to have an unconscious bias for ascribing the symptoms of coronary heart disease (CHD) among women to some other disorder [6]; and when the same proportion of women and men presented with chest pain, an observational study found that women were 2.5 times less likely to be referred to a cardiologist for management [7]. Similarly, it was shown that physicians tended to ask fewer diagnostic questions and prescribe the fewest CHD-related medications to middle-aged women [8]. In terms of ethnic bias, a systematic review of USA-based studies found that in the emergency room, black patients were 40% less likely to receive pain medication than white patients [9]. When such biases are present in training data or acquired during training, models have been found to perform unequally across different patient populations [10], and even negatively impact those in underrepresented groups [11]. For example, if a model was designed to determine who to prescribe CHD-related medications, men might be selected to receive the majority of them, further deepening inequities in healthcare.

For the purposes of our study, we developed two machine learning models that are unbiased towards different sensitive features – hospitals in different geographic regions and patients of different ethnicities. We focused on this problem in the context of rapid COVID-19 diagnosis; however, the methods described can be applied to many other scenarios where machine learning model are used to support decision-making. We compare our method to the benchmark set by XGBoost-based models [12], [13], and evaluate the generalizability of our models by performing prospective and external validation across emergency admissions to four independent UK National Health Service (NHS) Trusts.

### B. Related Works

The COVID-19 pandemic has highlighted the importance of data collaborations in order to rapidly respond to evolving and widespread global challenges. Recently, researchers showed that federated learning (FL) could effectively predict clinical outcomes of COVID-19, using combined data from multiple sites [14]. They found that FL could achieve high performance while maintaining data anonymity. Although FL can help remove some barriers to data sharing, it does not guarantee privacy, as model parameter updates can be used to infer sensitive information [15]. In our study, instead of focusing directly on privacy preservation, we focused on model fairness and bias mitigation. This is complementary to privacy preservation. For example, some locations/datasets may have very few patients of a given demographic, and thus, if a machine learning model is biased against this group, there is an increased probability of identifying these patients. To train fair and unbiased models, we applied a novel machine learning technique known as adversarial debiasing, where a model is trained to learn parameters that do not infer sensitive features. This technique has been used to develop models that output fair predictions, and has previously been successful in reducing gender (male versus female) bias in salary prediction [16], [17] and ethnicity (black vs white) bias in recidivism prediction [18]. There is currently no published research on the utility of adversarial debiasing in a clinical context. Additionally, all published adversarial debiasing research, thus far, has focused exclusively on debiasing binary attributes. However, in many real-world applications, it is often necessary to preserve a higher degree of granularity, as binning may not be biologically accurate and is heavily biased on the sample population. Therefore, through our study, we hope to encourage and demonstrate the effectiveness of adversarial debiasing on a wider range of prediction tasks and demographic features. To summarize, our main contributions in this paper are as follows:

- We propose a neural network-based framework based on adversarial debiasing that is capable of effectively determining COVID-19 status, while mitigating biases.
- We provide the first demonstration of adversarial debiasing in a clinical context, and evaluate its effectiveness across two different tasks - debiasing patient ethnicity and hospital location.
- We compare our results with the related previous work by Soltan et al. (2022) and perform external and prospective validation across four independent UK NHS hospital trusts, demonstrating the generalisability of our method.

## II. Methods

We trained neural network models to predict the COVID-19 status for patients attending hospital emergency departments (ED). Soltan et al. (2021) previously introduced an XGBoost-based machine learning pipeline for identifying patients with COVID-19 using blood tests, blood gas testing, and vital signs. They found that machine learning-based screening tests – CURIAL-1.0 [12] and CURIAL-Rapide/-Lab [13] – could rapidly detect COVID-19 amongst patients presenting to ED, and performed effectively as tests-of-exclusion (quick identification of patients who are most likely to test negative) during external validation across three NHS trusts. We aimed to build upon this existing work, developing the models with adversarial methods to effectively accomplish the same task, with the added capability of mitigating biases.

### A. Feature Set and Model Architecture

To train and validate our models, we used clinical data with linked, deidentified demographic information for patients across four hospital groups – Oxford University Hospitals NHS Foundation Trust (OUH), University Hospitals Birmingham NHS Trust (UHB), Bedfordshire Hospitals NHS Foundations Trust (BH), and Portsmouth Hospitals University NHS Trust (PUH). We performed prospective validation for patients presenting to OUH, and external validation for patients admitted to BH, PUH, and UHB.

For each of the models, a training set was used for model development, hyperparameter selection, training, and optimization; and after successful development and training, test sets were then used to evaluate the performance of the final models. How the data were split are detailed in following sections.

To better compare our results to the benchmark set by the CURIAL models, we used a similar set of features to CURIAL-Lab, which used a focused subset of routinely collected clinical features. These include blood tests (FBC, U&Es, liver function tests, CRP) and vital signs, excluding the coagulation panel and blood gas testing, which are not performed universally and are less informative [13]. However, unlike CURIAL-Lab, we did not include the type of oxygen delivery device as a feature. This was because it was not ranked as highly important for COVID-19 prediction, as determined by SHAP analysis [12] and, as neural networks evaluate features heavily on their variability with respect to other variables, we wanted to use a feature set consisting of entirely continuous variables to avoid potential optimization and convergence issues. Table I summarizes the final features included.

**TABLE I.**
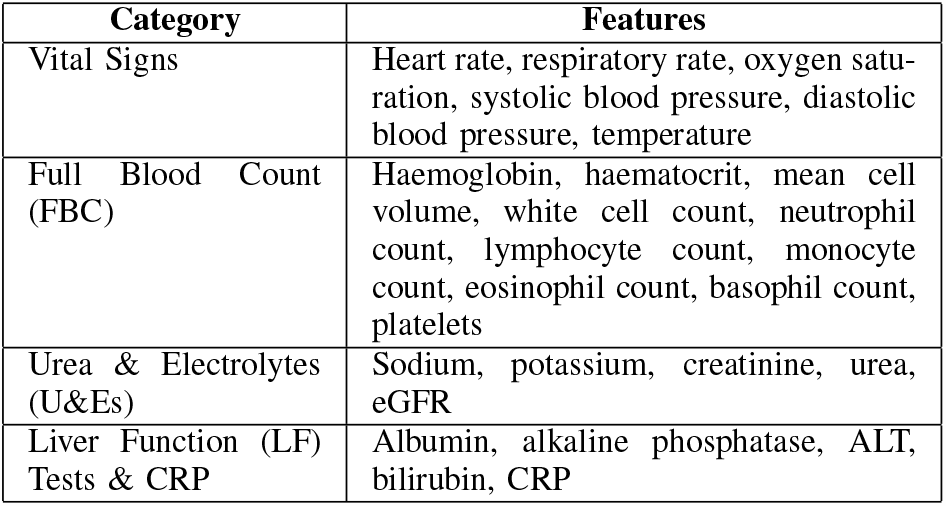
Clinical predictors considered. (ALT=alanine aminotransferase. CRP=C-reactive protein. eGFR=estimated glomerular filtration rate)

The adversarial debiasing architecture consists of two individual networks – a predictor network, *P*, and an adversary network, *A* (Fig. 1). *P* and *A* are each a multilayer perceptron (MLP) – the simplest form of a neural network. Here, *P* is trained to predict COVID-19 status, given a set of clinical features. Its raw output, *ŷ –* the predicted probability score, and the true label, *y*, are then used as the input to *A*, which tries to predict *z*. For our purposes, *z* is either hospital location or ethnicity (in machine learning literature, *z* is often referred to as the “protected feature”).

**Fig. 1.**
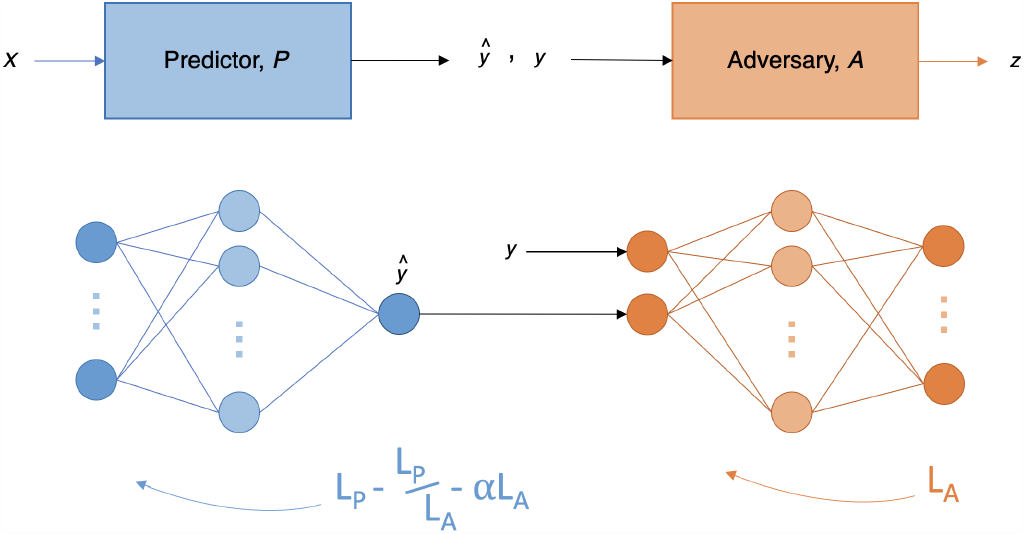
Adversarial training architecture.

Our goal is to train *P* to predict *y* effectively, regardless of the demographic membership of *z*. Thus, we want *P* to be able to accurately predict *y*, and *A* to poorly predict *z*, as this suggests that *P* has been trained in such a way that debiases *ŷ* with respect to *z*. We use cross-entropy loss (and binary cross-entropy loss when the feature is binary), where *L*_*P*_ represents the loss for *P*, and *L*_*A*_ represents the loss for *A*.

For *P* to be good at predicting *y* while being unbiased towards *z, P* is typically trained to balance the trade-off between the two losses. This is achieved using the combined loss function:

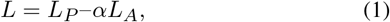

where *α* is an adjustable hyperparameter that signifies the importance of debiasing with respect to the protected feature, *z*. This combined function encourages *P* to minimize *L*_*P*_ while maximizing *L*_*A*_. However, to ensure that *P* never propagates in a direction that helps *A*, we modified the combined loss function to include a correction term. This is similar to the projection term previously introduced by Zhang et al. [17], which ensured that *P* never helped *A* decrease its loss. This projection term is the vector projection of the gradient of *L*_*P*_ onto the gradient of *L*_*A*_. Instead of modifying the gradients, we directly modified the loss function itself, such that the modified loss function for *P* becomes:

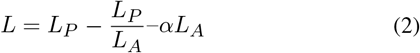

Under the assumption that *L*_*A*_ starts small (i.e. *A* is able to accurately predict *z*), the correction term, 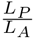, ensures that *P* propagates in the correct direction at the beginning of the training process.

For *P*, the sigmoid activation function is used in the output layer; and for *A*, the softmax activation function is used instead.

Using this framework, we developed two models: one for mitigating hospital location biases and one for mitigating patient ethnicity biases.

### B. Pre-Processing

Consistent with Soltan et al. [13], we addressed the presence of missing values by using population median imputation, then matched every positive COVID-19 presentation in the training set to a set of negative controls based on age, using a ratio of 20 controls:1 positive presentation.

We standardized all features in our data to have a mean of 0 and a standard deviation of 1. We also used the Synthetic Minority Oversampling Technique (SMOTE) [19] combined with Edited Nearest Neighbours sampling [20] to expand the minority class during training and remove any noisy or ambiguous samples, respectively. Implementation details can be found in Appendix I of the Supplementary Material.

### C. Hyperparameter Optimization

To estimate which hyperparameters would perform best for our task, we performed 5-fold cross-validation using the training data. We performed a grid search for different values of learning rate, number of hidden layer nodes in both the predictor and adversarial networks, and the dropout ratio. Details about the final hyperparameter values chosen for each model can be found in Appendix I of the Supplementary Material.

### D. Training Outline

The CURIAL models provided a benchmark to which we could compare our proposed models against. This allowed us to evaluate whether a neural network-based model (rather than a tree-based one) could effectively accomplish the main task, as well as ensure that we trained a strong classifier prior to the addition of any adversary component.

We began by training a predictor network without any adversary component. This was then used as a baseline to compare the relative effects of adversarial training. Thus, we trained a set of two models – a basic (standalone predictor) model and an adversarial model – for each of the protected attributes. Models were optimized to sensitivities of 0.9 to ensure that the model would be able to detect positive COVID-19 cases (this threshold was also used for the CURIAL models, allowing for direct comparison of results).

For the main task of predicting COVID-19 status in patients, we report sensitivity, specificity, positive and negative predictive values (PPV and NPV), and AUROC, alongside 95% confidence intervals (CIs) based on standard error. CIs for AUROC are calculated using Hanley and McNeil’s method [21]. Results are based on the evaluation of final, held-out test sets.

We evaluated the fairness of our models using the statistical definition of equalized odds, which states that a classifier is fair if true positive rates are equal and false positive rates are equal across all possible labels of the protected variable [22]. To assess multiple labels, we used the standard deviation (SD) of true positive and false positive scores. SD scores closer to zero suggest greater outcome fairness. The equations used to calculate true positive and false positive SD scores are as follows:

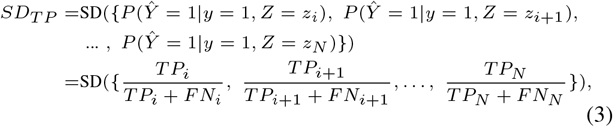

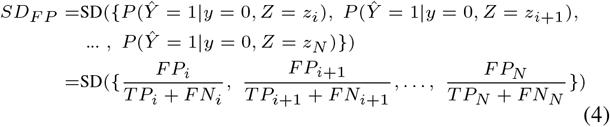

## III. Debiasing Ethnicity

### A. Patient Populations

For consistency, we trained our models using the same cohorts as those used to train and test CURIAL-Lab. We trained and optimized our model using 114,957 COVID-free patient presentations from OUH prior to the global COVID-19 outbreak, and 701 patient presentations during the first wave of the COVID-19 epidemic in the UK that had a positive PCR test for COVID-19. This ensured that the label of COVID-19 status was correct during training. We then validated the model on 72,223 admitted patients (4,600 COVID-19 positive with confirmatory testing) across four validation cohorts. A summary of each respective cohort is in Table II.

**TABLE II.** Summary population characteristics for OUH training cohorts, prospective validation cohort of patients attending OUH during the second wave of the UK COVID-19 epidemic, independent validation cohorts of patients admitted to three independent NHS Trusts. *indicates merging for statistical disclosure control.

|  | Training |  | Prospective validation | External Validation (Admissions) |  |  |
| --- | --- | --- | --- | --- | --- | --- |
|  | Oxford University Hospitals NHS Foundation Trust (pre-pandemic & wave 1 cases, to 30 June 2020) |  | Oxford University Hospitals NHS Foundation Trust | Portsmouth Hospitals NHS Trust | University Hospitals Birmingham NHS Foundation Trust | Bedfordshire Hospitals NHS Foundation Trust |
| Cohort | Pre-pandemic cohort | COVID-19-cases cohort | October 1, 2020 – March 6, 2021 | March 1, 2020 – February 28, 2021 | December 1, 2019 – October 29, 2020 | January 1, 2021 – March 31, 2021 |
| n, patients | 114,957 | 701 | 22,857 | 37,896 | 10,293 | 1,177 |
| n, COVID-19 PCR / genome test positive | 0 | 701 | 2,012 | 2,005 (5.29%) | 439 (4.27%) | 144 (12.2%) |
| Sex: |  |  |  |  |  |  |
| Male (%) | 53,370 (46.43) | 376 (53.64) | 11,409 (49.91) | 20,839 (54.99) | 4,831 (46.93) | 627 (53.27) |
| Female (%) | 61,587 (53.57) | 325 (46.36) | 11,448 (50.09) | 17,054 (45.0) | 5,462 (53.07) | 549 (46.64) |
| Age, yr (IQR) | 60 (38-76) | 72 (55-82) | 67 (49-80) | 69 (48-82) | 63 (42-79) | 68 (48-82) |
| Ethnicity: |  |  |  |  |  |  |
| White (%) | 93,921 (81.7) | 480 (68.47) | 17,387 (76.07) | 28,704 (75.74) | 6,848 (66.53) | 1,024 (87.0) |
| Not Stated (%) | 13,602 (11.83) | 128 (18.26) | 4,127 (18.06) | 8,389 (22.14) | 1,061 (10.31) | ≤ 10 |
| South Asian (%) | 2,754 (2.4) | 22 (3.14) | 441 (1.93) | 170 (0.45) | 1,357 (13.18) | 71 (6.03) |
| Chinese (%) | 284 (0.25) | * | 51 (0.22) | 42 (0.11) | 41 (0.4) | ≤ 10 |
| Black (%) | 1,418 (1.23) | 25 (3.57) | 279 (1.22) | 187 (0.49) | 484 (4.7) | 36 (3.06) |
| Other (%) | 1,840 (1.6) | 34 (4.85)* | 410 (1.79) | 269 (0.71) | 333 (3.24) | 29 (2.46) |
| Mixed (%) | 1,138 (0.99) | 12 (1.71) | 162 (0.71) | 135 (0.36) | 169 (1.64) | 13 (1.1) |

From Fig. 2, we can see that ethnicity is heavily skewed in our training dataset, making it a possible source of bias. Although “Unknown,” “Other,” and “Mixed” are ambiguous, we kept them in both our training and validation datasets, as they constituted a high number of total cases.

**Fig. 2.**
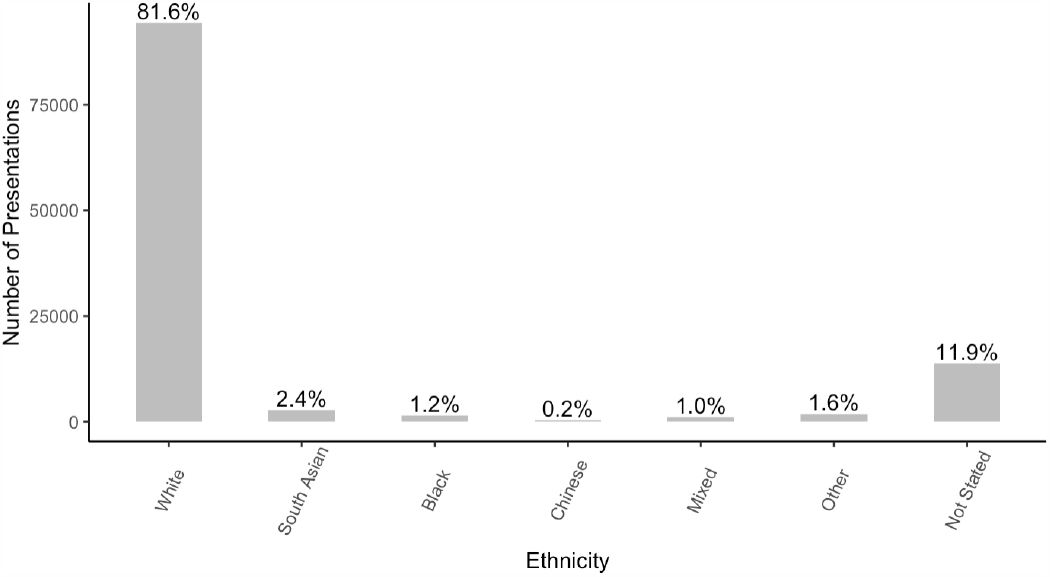
Distribution of patient ethnicities in OUH training cohort.

### B. Results

We prospectively and externally validated our models across four hospital cohorts. Using a sensitivity configuration of 0.9, AUROC scores for predicting COVID-19 status stayed consistent across both basic and adversarial models for each cohort, achieving the highest performance on the BH cohort (OUH: AUROC range 0.771-0.777 [CI range 0.759-0.789]; PUH: 0.744-0.765 [0.731-0.777]; UHB: 0.774-0.782 [0.748-0.808]; BH: 0.835-0.836 [0.793-0.878]). These were lower than the benchmark set by CURIAL-Lab (Soltan et al., 2021) (OUH: AUROC 0.878; PUH: 0.872 [CI 0.863 - 0.882]; UHB: 0.858 [0.838 - 0.878]; BH: 0.881 [0.851 - 0.912]). The optimized threshold also resulted in consistent scores for sensitivity across all models and cohorts (sensitivity range 0.844-0.868 with CI range 0.814-0.912).

And, consistent with CURIAL-Lab, our models also achieved high prevalence-dependent NPV scores (*>*0.98), demonstrating the ability for a neural network to exclude COVID-19 with high-confidence.

Although adversarial training only had a small effect on the overall performance of predicting COVID-19, relative to the basic model, it significantly changed the predicted probability outputs of the predictor in the adversarial model (Wilcoxon Signed Rank Test, p *<* 0.05 for all validation cohorts [Supplementary Table S5]).

For all validation cohorts, SD scores for true positive and false positive rates decreased when using an adversarial network (except for PUH, where the false positive rate remained the same). Overall, equalized odds were demonstrably improved through adversarial training, with only a slight trade-off in performance (AUROC decreased between 0.002-0.008 across OUH, PUH, and UHB cohorts). Complete performance and fairness metrics are shown in Fig. 3 and Table III (numerical results are shown in Supplementary Table S3).

**TABLE III.** Change in equalized odds during prospective validation and external validation for ethnicity-based adversarial training. Results reported as changes in SD of true positive and false positive rates after the addition of adversarial training.

|  | Prospective Validation |  | External Validation (Admissions) |  |  |  |  |  |
| --- | --- | --- | --- | --- | --- | --- | --- | --- |
| Cohort | Oxford University Hospitals |  | Portsmouth University Hospital NHS Trust |  | University Hospitals Birmingham NHS Foundation Trust |  | Bedfordshire Hospitals NHS Foundation Trust |  |
|  | n=22,857, prevalence=8.80% |  | n=37,896, prevalence=5.29% |  | n=10,293; prevalence=4.27% |  | n=1,177; prevalence=12.2% |  |
|  | Basic | Adv | Basic | Adv | Basic | Adv | Basic | Adv |
| True Positive Rate SD | 0.0387 | **0.0224** | 0.0490 | **0.0480** | 0.0985 | **0.0975** | 0.0964 | **0.0954** |
| False Positive Rate SD | 0.0574 | **0.0539** | 0.0361 | **0.0361** | 0.0510 | **0.0387** | 0.1039 | **0.0970** |

**Fig. 3.**
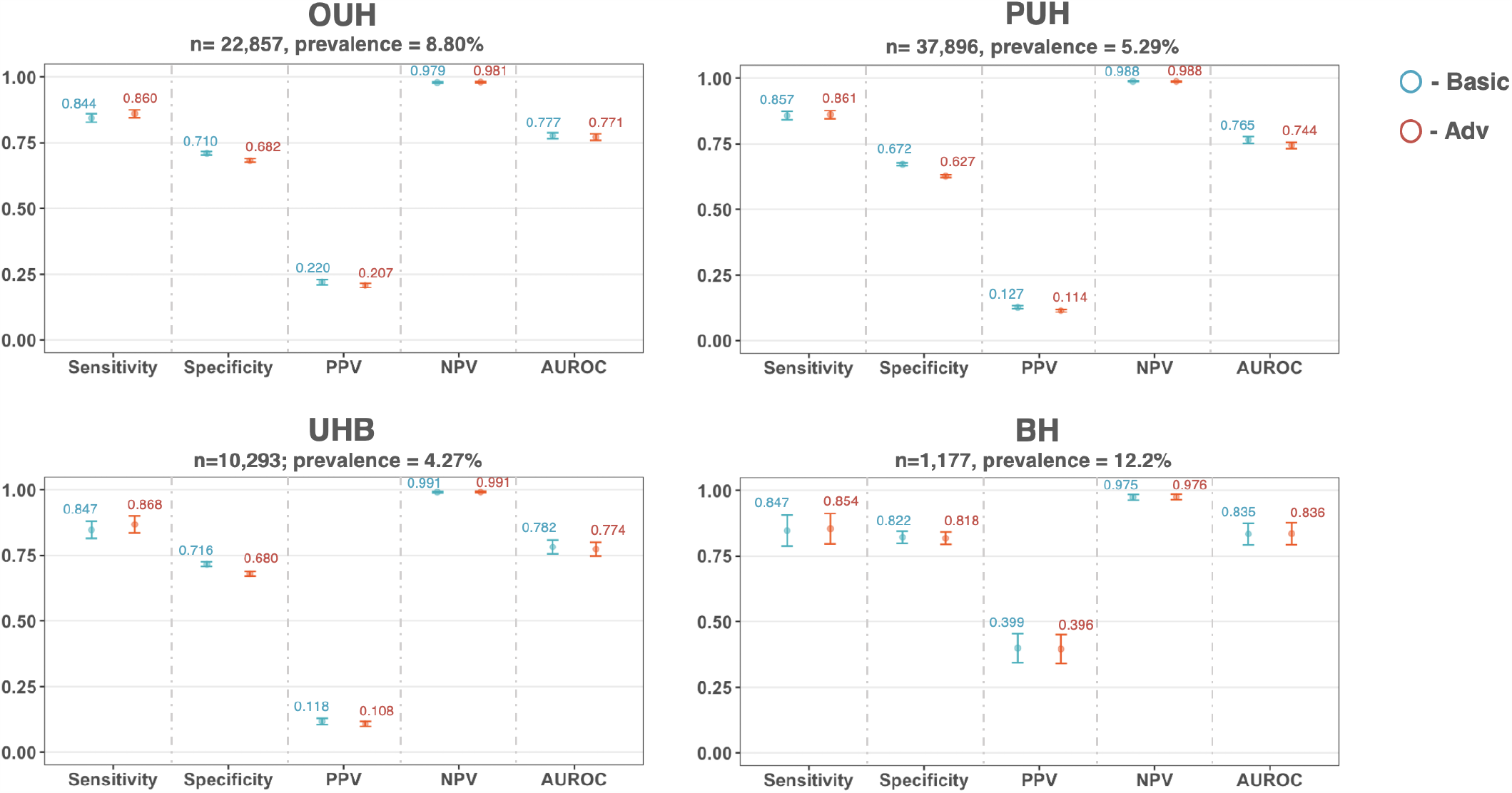
Performance of basic and adversarial models (blue and red, respectively) during prospective validation and external validation for ethnicity-based adversarial training. All models were optimized during training to achieve sensitivities of 0.9. Error bars show 95% confidence intervals. Numerical results are shown in Supplementary Table S3.

## IV. Debiasing Hospital Group

### A. Patient Populations

To further demonstrate the utility of our proposed method, we also trained a COVID-19 prediction model that is unbiased towards the hospital a patient attended. In order to evaluate bias related to hospital location, presentations from multiple sites needed to be present in the training data. Thus, we combined presentations from all hospital cohorts previously described (Table II), and used an 80:20 split to separate the data into training and test sets, respectively, stratified based on COVID-19 status and hospital cohort. This resulted in 150,304 presentations (4,249 COVID-19 positive) for training and optimization and 37,577 presentations (1,052 COVID-19 positive) for testing (Table IV).

**TABLE IV.** Training and test set distributions used for hospital-based adversarial training.

|  | Training | Test |
| --- | --- | --- |
| n, patients | 150,304 | 37,577 |
| n, COVID-19 positive | 4,249 (2.8%) | 1,052 (2.8%) |
| Hospital: OUH (%) | 110,906 (73.8%) | 27,609 (73.5%) |
| UHB (%) | 8,224 (5.5%) | 2,069 (5.5%) |
| BH (%) | 930 (0.6%) | 247 (0.7%) |
| PUH (%) | 30,244 (20.1%) | 7,652 (20.4%) |

As previously shown with ethnicity, we can see that the number of presentations available from different hospital cohorts is heavily skewed in our training dataset (Fig. 4), making it another possible source of bias.

**Fig. 4.**
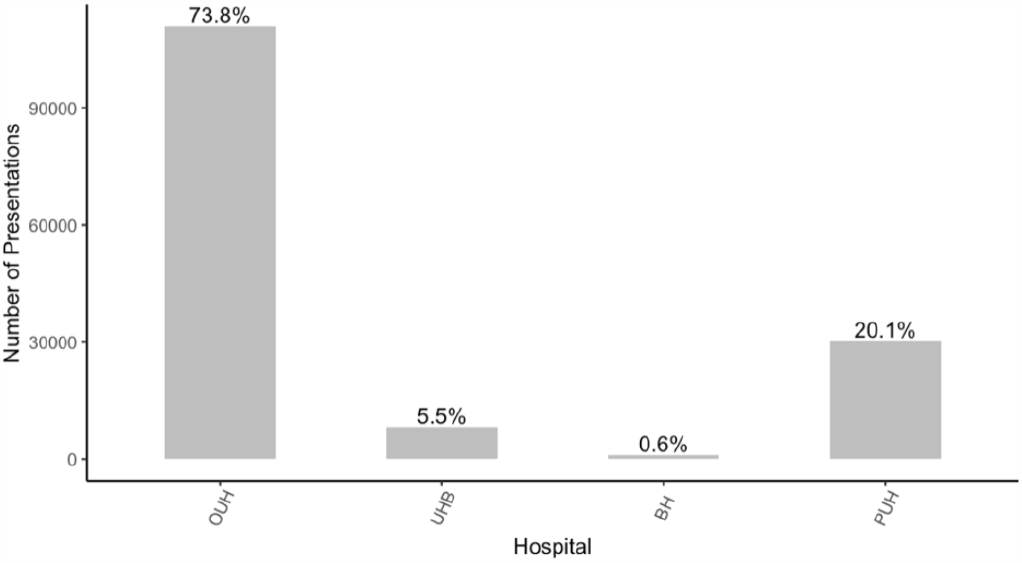
Distribution of hospital groups in training cohort.

Additionally, we used t-SNE to visualize a low-dimensional representation of all positive COVID-19 presentations in our training data (Fig. 5). From the results, we can see a distinct cluster (purple), which corresponds to a subset of the presentations from OUH. This suggests that the training data can be clustered by specific hospital groups (and thus, is biased), making this factor is an important and appropriate choice for bias mitigation.

**Fig. 5.**
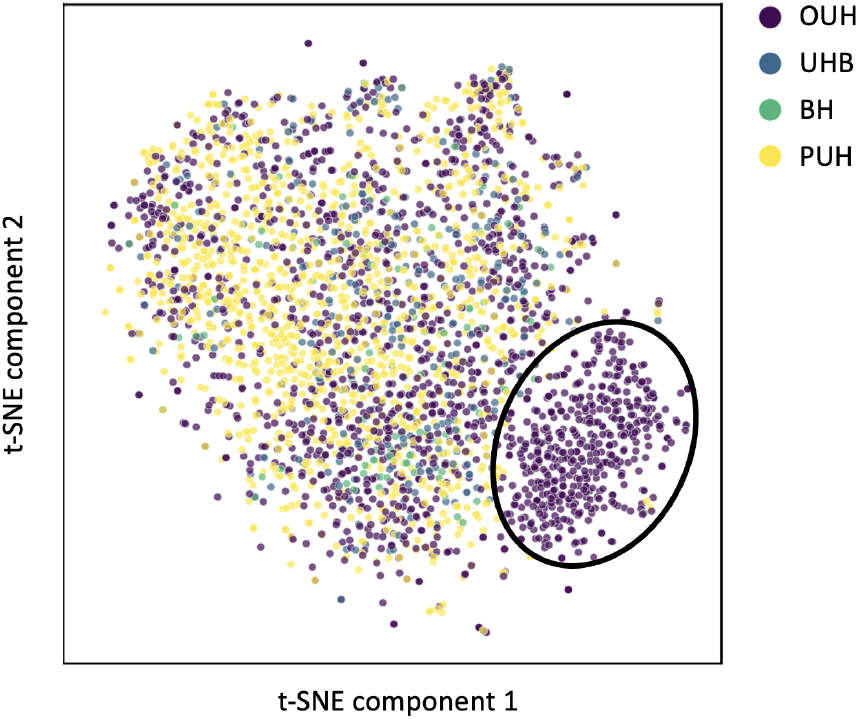
t-SNE plot of training data, labeled by hospital group.

### B. Results

We validated our models on a held-out set which included patient presentations from all four hospital cohorts. Using a sensitivity configuration of 0.9, model performance for predicting COVID-19 status was virtually the same for both the basic and adversarial models (AUROC 0.818 [CI 0.802-0.834]). This was lower than the benchmark set by CURIAL-Lab (Soltan et al., 2021) (AUROC range 0.842-0.868), however it was higher than the results achieved when attempting to mitigate ethnicity biases. Again, both models achieved high prevalence-dependent NPV scores (*>*0.99), demonstrating the ability for a neural network to exclude COVID-19 with high-confidence.

Relative to the basic model, adversarial training did not appear to affect the performance of predicting COVID-19 status. However, in terms of the output distribution, it significantly changed the predicted probability outputs between the two models. (Wilcoxon Signed Rank Test, p *<* 0.05 [Supplementary Table V]).

This affected the SD scores for both true positive and false positive rates, as both decreased with the addition of adversarial training. Thus, the adversarial model was able to improve equalized odds for hospital cohort, while maintaining its ability to perform the main task.

Complete performance and fairness metrics are shown in Fig. 6 and Table V (numerical results are shown in Supplementary Table S4).

**TABLE V.**
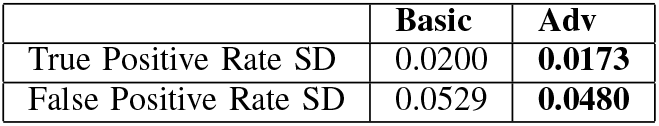
Change in equalized odds during validation for hospital-based adversarial training. Results reported as changes in SD of true positive and false positive rates after the addition of adversarial training.

|  | Basic | Adv |
| --- | --- | --- |
| True Positive Rate SD | 0.0200 | **0.0173** |
| False Positive Rate SD | 0.0529 | **0.0480** |

**Fig. 6.**
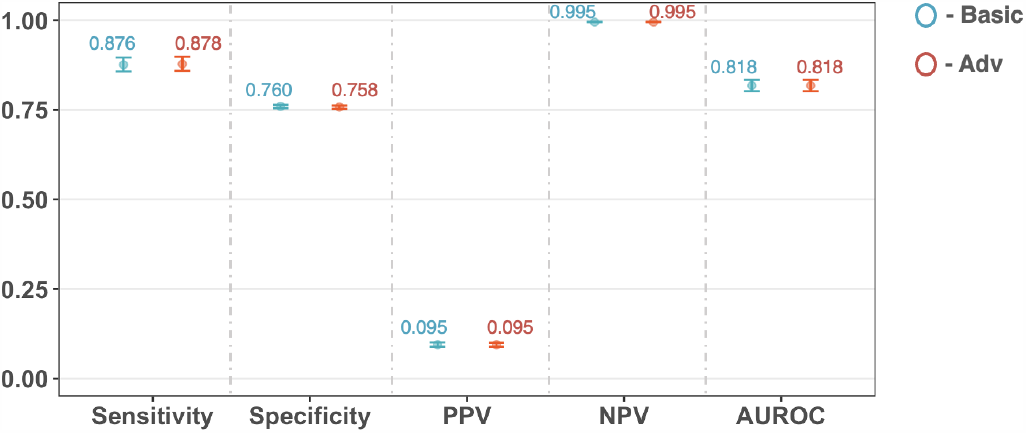
Performance of basic and adversarial models during prospective validation and external validation for hospital-based adversarial training. All models were optimized during training to achieve a sensitivity of 0.9. Error bars show 95% confidence intervals. Numerical results are shown in Supplementary Table S4.

## V. Discussion

The use of data-driven machine learning models is becoming increasingly prominent in healthcare settings. Thus, it is essential that these systems do not incorporate or reflect any discriminatory biases towards certain groups. In this study, we investigated a straightforward and effective method for training fair, unbiased machine learning models. We introduced an adversarial framework, and applied it to the main task of screening for COVID-19, while attempting to mitigate against hospital location and patient ethnicity biases. We also compared our method to the benchmark set by CURIAL-Lab.

Using the data visualization tool, t-SNE, we showed that variations between hospitals can be reflected by the data. These differences may be a result of different population distributions or lab analyzer brands used at each location. Thus, as machine learning datasets continue to expand, especially through collaborative efforts, greater attention needs to be given to bias mitigation, as different protocols and methods used to collect, process, and organize data can unintentionally encode biases.

We found that our basic neural network, trained independent of any adversary component, achieved slightly lower AUROCs compared to the benchmark set by CURIAL-Lab for ethnicity-based and hospital-based training (AUROC range 0.765-0.835 [CI range 0.828-0.873] and AUROC 0.818 [CI 0.802-0.834], respectively, versus AUROC range 0.858-0.881 [CI range 0.838-0.912]). However, this decrease in performance is reasonable, as neural networks can easily be overfit on training data and thus require very large amounts of balanced data to train, whereas XGBoost is known to be robust for rule-based classification tasks, even with smaller amounts of data present. This also explains why the basic network achieved better performance during the hospital-based bias mitigation task (compared to the ethnicity-based bias mitigation task), as a greater amount of combined data was used in training, further emphasizing the importance of collaborative approaches. In general, our neural network-based models still achieved high, clinically effective performances (NPVs *>* 0.98 at low prevalences). And, although AUROCs were slightly lower, a neural network-based model is advantageous in that it can be used in the adversarial framework outlined. This framework can be used with any model, regardless of complexity, as long as it is gradient descent-compatible. Thus, performance may be improved by alternative model architectures.

When an adversary network was trained against the main network, we found that the overall AUROCs achieved were consistent with those achieved by the basic model (average AUROC of 0.795 (0.030) and 0.790 (0.037) for basic and adversarial models, respectively). Additionally, in both cases, we found that the adversarial models were less biased than the basic one; thus, demonstrating that bias can be mitigated with minimal effect on performance. Although bias generally decreased, our models did not achieve complete equalized odds. One limitation could be that our data was skewed with respect to the protected features. As we are using neural networks, different distributions of the protected label can give significantly different results for the adversary model. This has previously been discussed [16], as using balanced data was found to have a much stronger effect on adversarial training. Thus, future experiments would greatly benefit from balanced training data.

With respect to debiasing against ethnicity, one limitation is the ambiguity of certain categories, namely, “Unknown,” “Mixed,” and “Other.” In our experiments, we kept these categories in order to maximize the number of cases (especially COVID-19 positive cases) used in training. This may have impacted the adversary network’s ability to confidently differentiate between different ethnicities, hindering its influence on the main network.

Bias may also still exist with respect to data missingness. Although we used population median imputation to “fill-in” missing values, the nature of the missing data may have conveyed important information, or reflected biases such as differences in access, practice, or recording protocols.

Another limitation is the difficulty in understanding how social, behavioral, and genetic factors independently and collectively impact outcomes. For example, consistent genetic effects across racial groups can result in genetic variants with a common biological effect; however, that effect can also be modified by both environmental exposures and the overall admixture of the population [23]. Thus, additional evaluations into the main prediction task (and related variables) will be necessary to determine what biases exist and how to best mitigate them.

## VI. Conclusion

In this study, we demonstrated that adversarial debiasing is a powerful technique for mitigating biases in machine learning models, providing the first piece of literature on adversarial debiasing in a clinical context. We trained our framework on a large, clinically-rich COVID-19 dataset, from four independent hospital cohorts, and demonstrated that the addition of an adversary model demonstrably improves outcome fairness, without compromising performance of the task at hand. We know that looking at variations across different regions and ethnic groups only addresses a small subset of existing inequities in healthcare; however, the framework we outlined can be easily applied to many different tasks and features. As technological capabilities continue to grow and machine learning continues to saturate decision-making processes in healthcare, we hope that the ability to develop fair models will encourage more hospitals to adopt machine learning-based technologies, and inspire greater confidence in the utility and reliability of these tools for making critical decisions.

## Data Availability

Data from OUH studied here are available from the Infections in Oxfordshire Research Database (https://oxfordbrc.nihr.ac.uk/research-themesoverview/antimicrobial- resistance-and-modernising-microbiology/infections-inoxfordshire- research-database-iord/), subject to an application meeting the ethical and governance requirements of the Database. Data from UHB, PUH and BH are available on reasonable request to the respective trusts, subject to HRA requirements. Code and supplementary information for this paper are available online alongside publication.

## Contributions

JY conceived the study, with design input from AS, YY & DAC. JY & AS preprocessed and verified the data. JY performed the analyses and wrote the manuscript. All authors revised the manuscript.

## Acknowledgements

We express our sincere thanks to all patients and staff across the four participating NHS trusts; Oxford University Hospitals NHS Foundation Trust, University Hospitals Birmingham NHS Trust, Bedfordshire Hospitals NHS Foundations Trust, and Portsmouth Hospitals University NHS Trust. We additionally express our gratitude to Jingyi Wang & Dr Jolene Atia at University Hospitals Birmingham NHS Foundation trust, Phillip Dickson at Bedfordshire Hospitals, and Paul Meredith at Portsmouth Hospitals University NHS Trust for assistance with data extraction.

## Ethics

United Kingdom National Health Service (NHS) approval via the national oversight/regulatory body, the Health Research Authority (HRA), has been granted for this work (IRAS ID: 281832).

## Data and Code Availability

Data from OUH studied here are available from the Infections in Oxfordshire Research Database (https://oxfordbrc.nihr.ac.uk/research-themesoverview/antimicrobial-resistance-and-modernising-microbiology/infections-inoxfordshire-research-database-iord/), subject to an application meeting the ethical and governance requirements of the Database. Data from UHB, PUH and BH are available on reasonable request to the respective trusts, subject to HRA requirements. Code and supplementary information for this paper are available online alongside publication.

