## Supplementary Material for "Algorithmic Fairness and Bias Mitigation for Clinical Machine Learning: Insights from Rapid COVID-19 Diagnosis by Adversarial Learning"

### Appendix I
### Software Packages, Implementation, and Hyperparameter Values

Models were implemented using Python (v3.6.9) and PyTorch (v1.7.0). Scikit Learn (v0.24.1) was used for standardization, median imputation, and calculating performance metrics. Imbalanced Learn (v0.7.0) was used to implement SMOTE and Edited Nearest Neighbors Sampling. Performance metrics were calculated using Scikit Learn and fairness metrics were manually programmed. t-SNE was implemented using Scikit Learn, with a perplexity of 40 and early exaggeration of 30.

TABLE S1
Ethnicity-based Adversarial Training Final Hyperparameters

| **Learning Rate** | **N_p** | **N_adv** | **Dropout** | **Alpha** | **Epochs** | **Optimizer** |
|---|---|---|---|---|---|---|
| 1e-4 | 10 | 10 | 0.3 | 10 | 4000 | Adam |

TABLE S2
Hospital-based Adversarial Training Final Hyperparameters

| **Learning Rate** | **N_p** | **N_adv** | **Dropout** | **Alpha** | **Epochs** | **Optimizer** |
|---|---|---|---|---|---|---|
| 1e-4 | 100 | 10 | 0.3 | 1 | 4000 | Adam |

### Appendix II
### Ethnicity-based Adversarial Training Results

TABLE S3
COVID-19 Prediction Performance

| | **Prospective Validation** | | **External Validation (Admissions)** | | | | | |
|---|---|---|---|---|---|---|---|---|
| Cohort | Oxford University Hospitals | | Portsmouth University Hospital NHS Trust | | University Hospitals Birmingham NHS Foundation Trust | | Bedfordshire Hospitals NHS Foundation Trust | |
| | n= 22,857, prevalence = 8.80% | | n= 37,896, prevalence = 5.29% | | n=10,293, prevalence = 4.27% | | n=1,177, prevalence = 12.2% | |
| | Basic | Adv | Basic | Adv | Basic | Adv | Basic | Adv |
| Sensitivity | 0.844 (0.828-0.860) | 0.860 (0.845-0.875) | 0.857 (0.842-0.873) | 0.861 (0.846-0.876) | 0.847 (0.814-0.881) | 0.868 (0.836-0.900) | 0.847 (0.789-0.906) | 0.854 (0.797-0.912) |
| Specificity | 0.710 (0.704-0.717) | 0.682 (0.676-0.689) | 0.672 (0.667-0.677) | 0.627 (0.622-0.632) | 0.716 (0.708-0.725) | 0.680 (0.671-0.690) | 0.822 (0.799-0.845) | 0.818 (0.795-0.842) |
| PPV | 0.220 (0.210-0.229) | 0.207 (0.199-0.216) | 0.127 (0.122-0.133) | 0.114 (0.109-0.119) | 0.118 (0.106-0.129) | 0.108 (0.098-0.118) | 0.399 (0.344-0.454) | 0.396 (0.341-0.450) |
| NPV | 0.979 (0.977-0.982) | 0.981 (0.978-0.983) | 0.988 (0.987-0.990) | 0.988 (0.986-0.989) | 0.991 (0.988-0.993) | 0.991 (0.989-0.994) | 0.975 (0.964-0.985) | 0.976 (0.966-0.986) |
| F1 | 0.348 | 0.334 | 0.222 | 0.202 | 0.206 | 0.192 | 0.542 | 0.541 |
| AUROC | 0.777 (0.765-0.789) | 0.771 (0.759-0.784) | 0.765 (0.752-0.777) | 0.744 (0.731-0.756) | 0.782 (0.756-0.808) | 0.774 (0.748-0.800) | 0.835 (0.793-0.876) | 0.836 (0.794-0.878) |

Performance of basic and adversarial models during prospective validation and external validation for ethnicity-based adversarial training. All models were optimized during training to achieve sensitivities of 0.9. Results are reported alongside 95% confidence intervals.

### APPENDIX III
### HOSPITAL-BASED ADVERSARIAL TRAINING RESULTS

TABLE S4
COVID-19 Prediction Performance

| | **Basic** | **Adv** |
|---|---|---|
| Sensitivity (%) | 0.876 (0.857-0.896) | 0.878 (0.859-0.898) |
| Specificity (%) | 0.760 (0.755-0.764) | 0.758 (0.753-0.762) |
| PPV (%) | 0.095 (0.089-0.101) | 0.095 (0.089-0.100) |
| NPV (%) | 0.995 (0.995-0.996) | 0.995 (0.995-0.996) |
| F1 | 0.171 | 0.171 |
| AUROC | 0.818 (0.802-0.834) | 0.818 (0.802-0.834) |

Performance of basic and adversarial models during prospective validation and external validation for hospital-based adversarial training. All models were optimized during training to achieve sensitivities of 0.9. Results are reported alongside 95% confidence intervals.

### APPENDIX IV
### BASIC AND ADVERSARIAL MODEL COMPARISON

TABLE S5
Comparing Outputs of Basic and Adversarial Models (p-values)

<table>
<tr><th colspan="4">Debiasing Ethnicity</th><th>Debiasing Hospital</th></tr>
<tr><td>Prospective Validation</td><td colspan="3">External Validation</td><td rowspan="2">Validation Set</td></tr>
<tr><td>OUH</td><td>PUH</td><td>UHB</td><td>BH</td></tr>
<tr><td><e-308</td><td><e-308</td><td><e-308</td><td>8.217e-83</td><td><e-308</td></tr>
</table>

Wilcoxon Signed Rank Test implemented using the statistics package from the SciPy library (scipy.stats). Numbers smaller than <e-308 cannot be distinguished from 0 by floating-point types in Python.
